# Antihypertensive medication uses and serum ACE2 levels

**DOI:** 10.1101/2020.05.21.20108738

**Authors:** Valur Emilsson, Elias F. Gudmundsson, Thor Aspelund, Brynjolfur G. Jonsson, Alexander Gudjonsson, Lenore J. Launer, Lori L. Jennings, Valborg Gudmundsdottir, Vilmundur Gudnason

**Affiliations:** Icelandic Heart Association, Holtasmari 1, IS-201 Kopavogur, Iceland; Faculty of Medicine, University of Iceland, 101 Reykjavik, Iceland; Laboratory of Epidemiology and Population Sciences, Intramural Research Program, National Institute on Aging, Bethesda, MD 20892-9205, USA; Novartis Institutes for Biomedical Research, 22 Windsor Street, Cambridge, MA 02139, USA

**Author notes:** Corresponding authors: Valur Emilsson and Vilmundur Gudnason, Icelandic Heart Association, Holtasmari 1, IS-201 Kopavogur, Iceland.

## Abstract

**Importance:** Recent reports have shown that hypertension is the most common comorbidity associated with mortality in the current coronavirus disease 2019 (COVID-19). This has been related to the use of angiotensin-converting enzyme inhibitors (ACEIs) and angiotensin II receptor blockers (ARBs) as animal studies indicate that these medications increase levels of ACE2, the cellular entry point for the coronavirus SARS-CoV-2. This has prompted clinicians to recommend discontinuing ACEIs and ARBs.

**Objective:** To examine the effect of ACEIs or ARBs treatment on serum levels of ACE2 and other key enzymes in the renin-angiotensin system (RAS).

**Design, Setting, and Participants:** A single center population-based study of 5457 Icelanders from the Age, Gene/Environment Susceptibility Reykjavik Study (AGES-RS) of the elderly (mean age 75±6 years) stratified by ACEIs (N = 699) or ARBs (N = 753) treatment.

**Main Outcomes and Measures:** The AGES-RS study population was stratified by ACEIs and ARBs medication use and compared for age, body mass index (BMI) (kg/m^2^), hypertension and type 2 diabetes (T2D) as well as serum levels of renin, ACE and ACE2.

**Results:** While renin and ACE levels were significantly raised in serum of individuals on ACEIs or ARBs treatments, the ACE2 levels remained unaffected.

**Conclusions and Relevance:** Treatment with ACEIs or ARBs does not raise ACE2 levels in serum. Therefore, the present study does not support the proposed discontinuation of these medications among patients affected with COVID-19.

**Key Points:** *Question:* Does treatment with the antihypertensive medications angiotensin-converting enzyme inhibitors (ACEIs) and angiotensin II receptor blockers (ARBs) result in elevated levels of the cellular receptor for the coronavirus SARS-CoV-2, ACE2?

*Findings:* In a single center population-based cohort (AGES-RS), 699 and 753 individuals were either on ACEIs or ARBs treatment, respectively. The serum levels of the key enzymes in the renin-angiotensin system (RAS), renin, ACE and ACE2 were measured in 5457 subjects of the AGES-RS and their serum levels in individuals on ACEIs or ARBs treatment compared to those not using these medications. While renin and ACE were significantly raised in serum of ACEIs and ARBs users, the levels of ACE2 remained unaffected.

*Meaning:* These results do not support the proposed routine discontinuation of ACEIs or ARBs among patients affected with COVID-19.

## Introduction

The current coronavirus disease 2019 (COVID-19) pandemic, caused by the severe acute respiratory syndrome coronavirus 2 (SARS-CoV-2), is associated with major respiratory failure where the old and those with an underlying chronic disease are at highest risk of mortality^1^. The most frequent comorbidities associated with COVID-19 related mortality are hypertension and type 2 diabetes (T2D)^2,3^. Although lower survival can simply be attributed to the frailty of this population, it has been suggested that administration of angiotensin-converting enzyme inhibitors (ACEIs) and angiotensin receptor blockers (ARBs) may affect the susceptibility to COVID-19 related outcomes by upregulating ACE2^4^ It is well known that ACE2 is the cellular receptor that SARS-CoV-2 and other SARS coronaviruses bind to for entering the host cell^5^.

The evidence for ACEIs and ARBs upregulating ACE2 levels comes predominantly from animal studies of the heart tissue^6^, while a similar effect has not been established in pulmonary tissues or serum. In this case of upregulation, it is possible that the finding of a higher mortality in hypertensive patients is confounded by levels of ACE2 raised in those using ACEIs or ARBs. This has prompted clinicians to recommend discontinuing ACEIs and ARBs when patients are treated for COVID-19^7^. This claim has been debated but there is inadequate clinical and scientific evidence for the above-mentioned connection. Further, any decision to discontinue those medications may put patients at risk, especially the elderly. No large population-based cohort studies containing measurements of ACE2 levels and different antihypertensive treatment groups have been published to date that address potential effect of ACEIs/ARBs medications on ACE2 levels. In the present study, we examined the effect of ACEIs and ARBs use on the serum levels of the key enzymes in the circulatory renin-angiotensin system (RAS), renin, ACE and ACE2 in a population of 5457 individuals aged 65 and above.

## Methods

### Study population

Participants aged 66 through 96 were from the Age, Gene/Environment Susceptibility Reykjavik Study (AGES-RS) cohort^8^, a single-center prospective population-based study aimed to understand aging in the context of gene environment interactions. Descriptive statistics of this cohort as well as detailed definition of the various disease end-points and relevant phenotypes measured have been published elsewhere^8,9^. The AGES-RS was approved by the NBC in Iceland (approval number VSN-00-063), and by the National Institute on Aging Intramural Institutional Review Board (U.S.) and the Data Protection Authority in Iceland.

### Study design and statistical analysis

Baseline characteristics for the AGES-RS study population were compared by treatments of ACEIs, ARBs or either medication group (**Table 1**). Means and standard deviations were presented for continuous variables and numbers and percentages for categorical variables. Differences between treatment groups were tested using two tailed T-tests for continuous variables and χ^2^ tests for categorical variables. Treatment group differences were compared for the following characteristics: age, body mass index (BMI) (kg/m^2^), obesity defined as BMI>30, measured hypertension defined as systolic blood pressure >140 or diastolic blood pressure >90, any indication of hypertension defined as having high blood pressure or taking any blood pressure medication or by questionnaire. Type 2 diabetes was defined as fasting serum glucose 126 mg/dL (7.0 mmol/L) or self-reported history of diabetes or the use of insulin or oral glucose-lowering medications. Participants brought all medications used during the two weeks prior to recruitment and the use of ACEIs and ARBs recorded according to ATC codes.

**Table 1:**
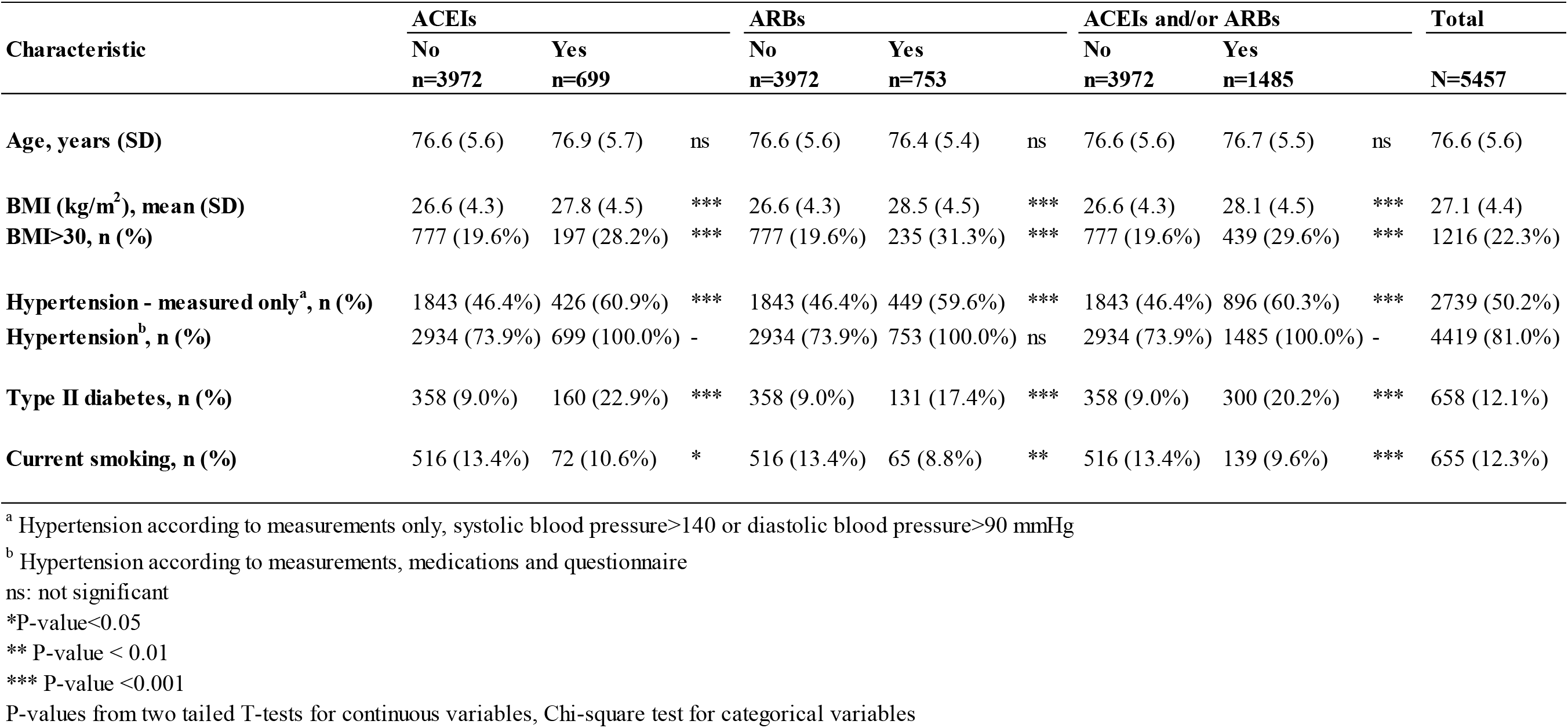
Description of the AGES study population (N = 5457, 57% females), stratified on ACEIs and ARBs use

### Protein measurements

Serum protein levels of renin, ACE and ACE2 were measured in 5457 individuals from the AGES-RS using the aptamer-based protein profiling platform^9^. Various metrics related to the performance of the proteomic platform including aptamer specificity, assay variability and reproducibility have already been described^9^. Proteins were investigated on a log2 scale where one-unit increase represents a doubling in protein levels. Observed levels of ACE2, ACE and renin were graphically presented by ACEIs and ARBs treatments using violin plots. General linear regression models were used to estimate differences in protein levels between treatment groups with confidence intervals and P-values. Adjustment in the models was made for the covariates age and sex. A P-value <0.05 was considered statistically significant for all hypothesis testing.

## Results

In the present study we examined the effect of ACEIs and ARBs treatment on ACE2 serum levels in a population of 5457 individuals from the deeply phenotyped AGES-RS^8^. The sera from each AGES-RS participant was measured for 4782 proteins, including renin, ACE and ACE2, using the aptamer-based multiplex proteomic technology^9^. A total of 1485 individuals were on either ACEIs or ARBs treatment (**Table 1**), of which 33 were on both medications. As expected, the individuals using antihypertensive drugs were more likely to manifest with hypertension, obesity and/or T2D (**Table 1**). The violin plots in **Figure 1** demonstrate the levels (log2 scale) of renin, ACE and ACE2 in serum from individuals that are either on ACEIs or ARBs treatment or no treatment. Treatment with ACEIs or ARBs was not associated with higher serum ACE2 levels (**Figure 1**). In fact, we observed a weak trend of lower ACE2 levels in individuals on ARBs treatment (β = -0.0222, P = 0.043) while ACEIs showed no effect on ACE2 (β = -0.0003, P = 0.976). The increase observed in ACE levels with ACEIs treatment (β = 0.2899, P < 0.001) is consistent with available data showing positive effect of ACEIs use on ACE mRNA levels and activity^10^. In contrast, treatment with ARBs had no effect on ACE levels (β = -0.0005, P = 0.699). As was expected, serum renin levels were elevated in response to both ACEIs (β = 0. 4994, P < 0.001) and ARBs (β = 0.5243, P < 0.001) treatment, as these medications diminish the negative feedback regulation of angiotensin II on renal secretion^11^. In summary, our results do not support the hypothesis that treatment with ACEIs or ARBs upregulates ACE2 as previously noted in animal studies.

**Figure 1.**
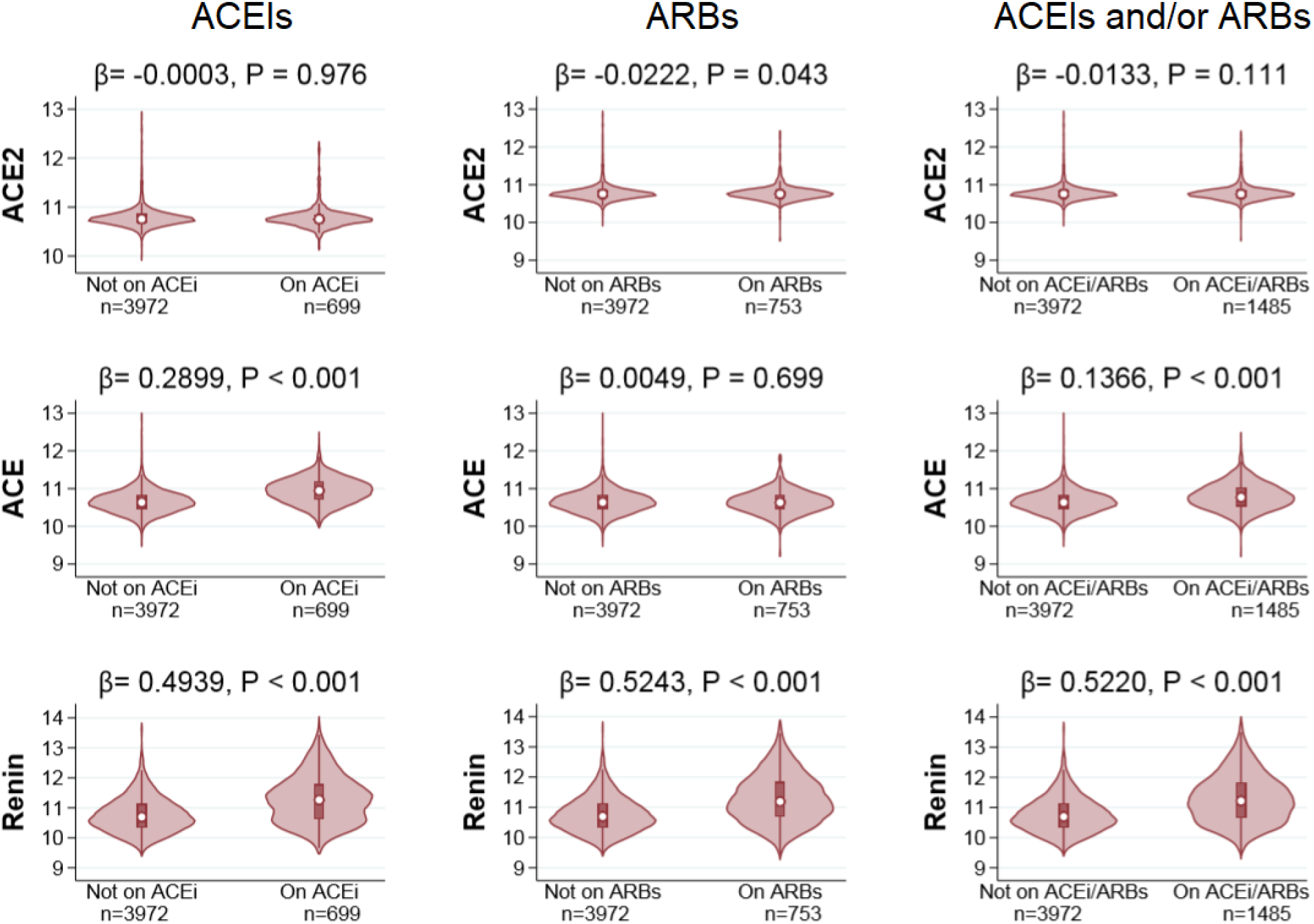
Observed levels of ACE2, ACE and renin in response to treatment of ACEIs (left panel), ARBs (middle panel) and ACEIs or ARBs (right panel). The y-axis represents log2-transformed distribution of each protein. General linear regression models were used to estimate differences in protein levels between treatment groups with confidence intervals and P-values. Adjustment in the models was made for the covariates age and sex. We note that the outliers have been removed.

## Discussion

The circulatory RAS is an important hormonal pathway in the development of hypertension where the enzymes renin, ACE and ACE2 play key regulatory roles^11^. Unlike renin, which is secreted, the ACE and ACE2 are membrane-bound ectoenzymes that are released into the circulation *via* ectodomain shedding^12^. Variable levels of these enzymes in serum are likely a result of genetic factors, differential gene expression and/or ectodomain shedding, often influenced by disease or administration of drugs *via* negative or positive feedback loops of the renin-angiotensin system^13^. Genetic variants affecting ACE mRNA and protein levels in lung tissues and plasma, respectively, have recently been used as proxies to investigate the potential influence of ACEIs on ACE2 levels^14^. Here, low expresser ACE alleles that are supposed to mirror ACEI treatment were not associated with changes in ACE2 levels.

Proteins in circulation emanate from virtually every tissue of the body^9^. Thus, we cannot easily determine how much each tissue contributes to ACE2 release into blood. For instance, it is possible that the effect of ACEIs and ARBs on ACE2 levels in pulmonary tissues manifests differently from what was observed in serum. Moreover, just how the balance between the levels of soluble ACE2 and its membrane-bound receptor counterpart is maintained, has still to be fully defined. However, our results using a large population study with many on ACEIs and ARBs treatment agree well with a study showing genetic proxies for ACE inhibition having no effect on ACE2 levels in pulmonary tissues and in plasma^14^, as well as a recent report on a lack of association between ACEIs/ARBs use and severity of outcomes in COVID-19 among 115 hospitalized male patients^15^. The accumulated data do not back the notion that the use of the antihypertensive medications ACEIs and ARBs results in elevation of ACE2, that in turn could lead to increased susceptibility to severe outcome in COVID-19. Importantly, these results do not support the proposed routine discontinuation of ACEIs or ARBs among patients affected with COVID-19.

## Limitations

The present study included many individuals that were on ACEIs or ARBs treatment, thus providing adequate power to assess potential effect of these medications on ACE2 levels in serum. However, the lack of association of ACEIs/ARBs to variable ACE2 levels in the serum warrants further validation in other study populations. Furthermore, the results presented may be limited to that of plasma and serum and may not reflect the effect of these medications on ACE2 levels in solid tissues like pulmonary tissues. Also, the measured circulating levels of ACE2 may not reflect its cell surface receptor levels in solid tissues. Finally, all participants of the AGES-RS are white (Caucasians) which may limit the transferability and generalizability of the results as ACE2 levels differ across races and ethnicities^16^.

## Conclusions

The ACEIs and ARBs are common medications used to treat hypertension. Withdrawal of these medications can put people at risk of developing serious outcomes. In this study, we did not find evidence for use of ACEIs and ARBs leading to raised ACE2 levels, at least not in serum, that in turn could lead to increased susceptibility to severe outcome in COVID-19. These results do not support the proposed discontinuation of ACEIs or ARBs for patients with COVID-19.

## Data Availability

The custom-design Novartis SOMAscan is available through a collaboration agreement with the Novartis Institutes for BioMedical Research. Data from the AGES Reykjavik study are available through collaboration under a data usage agreement with the IHA.

## Acknowledgment

Supported by Novartis Institute for Biomedical Research (NIBR), Icelandic Heart Association, and in part by the intramural research program at the National Institute of Aging (N01-AG-12100 and HHSN271201200022C). We thank all AGES-Reykjavik study participants, and the staff of the IHA for their contribution to the AGES-Reykjavik study. The custom-design Novartis SOMAscan is available through a collaboration agreement with the Novartis Institutes for BioMedical Research. Data from the AGES Reykjavik study are available through collaboration under a data usage agreement with the IHA. L.L.J. is an employee of and own stocks in Novartis. All other authors declare they have no competing interests.

